# Plasma Proteome Variation and its Genetic Determinants in Children and Adolescents

**DOI:** 10.1101/2023.03.31.23287853

**Authors:** Lili Niu, Sara Elizabeth Stinson, Louise Aas Holm, Morten Asp Vonsild Lund, Cilius Esmann Fonvig, Leonardo Cobuccio, Jonas Meisner, Helene Bæk Juel, Maja Thiele, Aleksander Krag, Jens-Christian Holm, Simon Rasmussen, Torben Hansen, Matthias Mann

**Affiliations:** Novo Nordisk Foundation Center for Protein Research, University of Copenhagen, DK-2200 Copenhagen, Denmark; Department of Proteomics and Signal Transduction, Max Planck Institute of Biochemistry, 82152 Martinsried, Germany; Novo Nordisk Foundation Center for Basic Metabolic Research, University of Copenhagen, DK-2200 Copenhagen, Denmark; The Children’s Obesity Clinic, accredited European Centre for Obesity Management, Department of Pediatrics, Copenhagen University Hospital Holbæk, DK-4300 Holbæk, Denmark; Department of Biomedical Sciences, University of Copenhagen, DK-2200 Copenhagen, Denmark; The Faculty of Health and Medical Sciences, University of Copenhagen, DK-2200 Copenhagen, Denmark; Odense Liver Research Centre, Department of Gastroenterology and Hepatology, Odense University Hospital, DK-5000 Odense, Denmark; Department of Clinical Research, University of Southern Denmark, DK-5000 Odense, Denmark; The Novo Nordisk Foundation Center for Genomic Mechanisms of Disease, Broad Institute of MIT and Harvard, Cambridge, MA 02142, USA

## Abstract

The levels of specific proteins in human blood are the most commonly used indicators of potential health-related problems^1^. Understanding the genetic and other determinants of the human plasma proteome can aid in biomarker research and drug development. Diverse factors including genetics, age, sex, body mass index (BMI), growth and development including puberty can affect the circulating levels of proteins^2–5^. Affinity-based proteomics can infer the relationship between blood protein levels and these factors at a large scale^6–10^. Compared to these methods, mass spectrometry (MS)-based proteomics provides much higher specificity of identification and quantification^11–13^, but existing studies are limited by small sample sizes or low numbers of quantified proteins^14–17^. Here we aim to elucidate to which extent genomic variation affects plasma protein levels across diverse age ranges and cohort characteristics. Employing a streamlined and highly quantitative MS-based plasma proteomics workflow, we measured the plasma proteome of 2,147 children and adolescents. Levels of 90% of these proteins were significantly associated with age, sex, BMI or genetics. More than 1,000 protein quantitative trait loci (pQTLs) – a third of which were novel – regulated protein levels between a few percent and up to 30-fold. These replicated excellently in an independent cohort of 558 adults, with highly concordant effect sizes (Pearson’s r > 0.97). We developed a framework to eliminate artefactual pQTLs due to protein-altering variants, paving the way for large-scale interrogation of pQTLs using MS-based proteomics. Our data reveal unexpectedly extensive genetic impacts on plasma protein levels, consistent from childhood into adulthood. These findings have implications for biomarker research and drug development.

**Highlights:**

1. First large-scale proteome-wide and genome-wide association study in children and adolescents
2. MS-based proteomics achieves very high specificity and quantitative accuracy
3. Robust plasma protein trajectories during development predict age and body mass index
4. Largest set of pQTLs for plasma proteome by MS-based proteomics
5. pQTLs are highly replicable between children and adults
6. Large-scale pQTL identification enables generic drug target validation

## Discovery and replication cohorts

Our cohort of 2,147 children and adolescents aged 5-20 years for discovery consists of individuals from the HOLBAEK Study including the general population (45%) and the Children’s Obesity Clinic in Holbæk, Denmark (55%) (Fig. 1a). For replication of the genetic effects on plasma protein levels, we used a cohort of adults (n=558) with alcohol-related liver disease (ALD) aged 19-82 years matched with healthy controls recruited from the Region of Southern Denmark^18^. Supplementary Table 1 provides baseline participant characteristics for both cohorts, on which we conducted single nucleotide polymorphism (SNP)-based genotyping and MS-based plasma proteome profiling (Fig. 1b and Methods). We acquired plasma proteome profiles of all participants (n□=□2,147) with a data-independent acquisition (DIA) strategy^19^ and a single-run workflow using a liquid chromatography system designed for robust clinical use^20^ coupled online to a high resolution Orbitrap mass spectrometer^21^. After stringent quality control, the filtered dataset contained 420 proteins with a data completeness of 88%. Analyzing 94 quality assessment samples over a six-week measurement period revealed an 18% median coefficient of variation for the entire workflow (Extended Data Fig. 1). We estimated the effect of age, sex, BMI SDS (BMI standard deviation score adjusting for age and sex)^22^, and 5.2 million SNPs on the quantitative levels of these plasma proteins using a high-performance computing server (Fig. 1d).

**Fig. 1.**
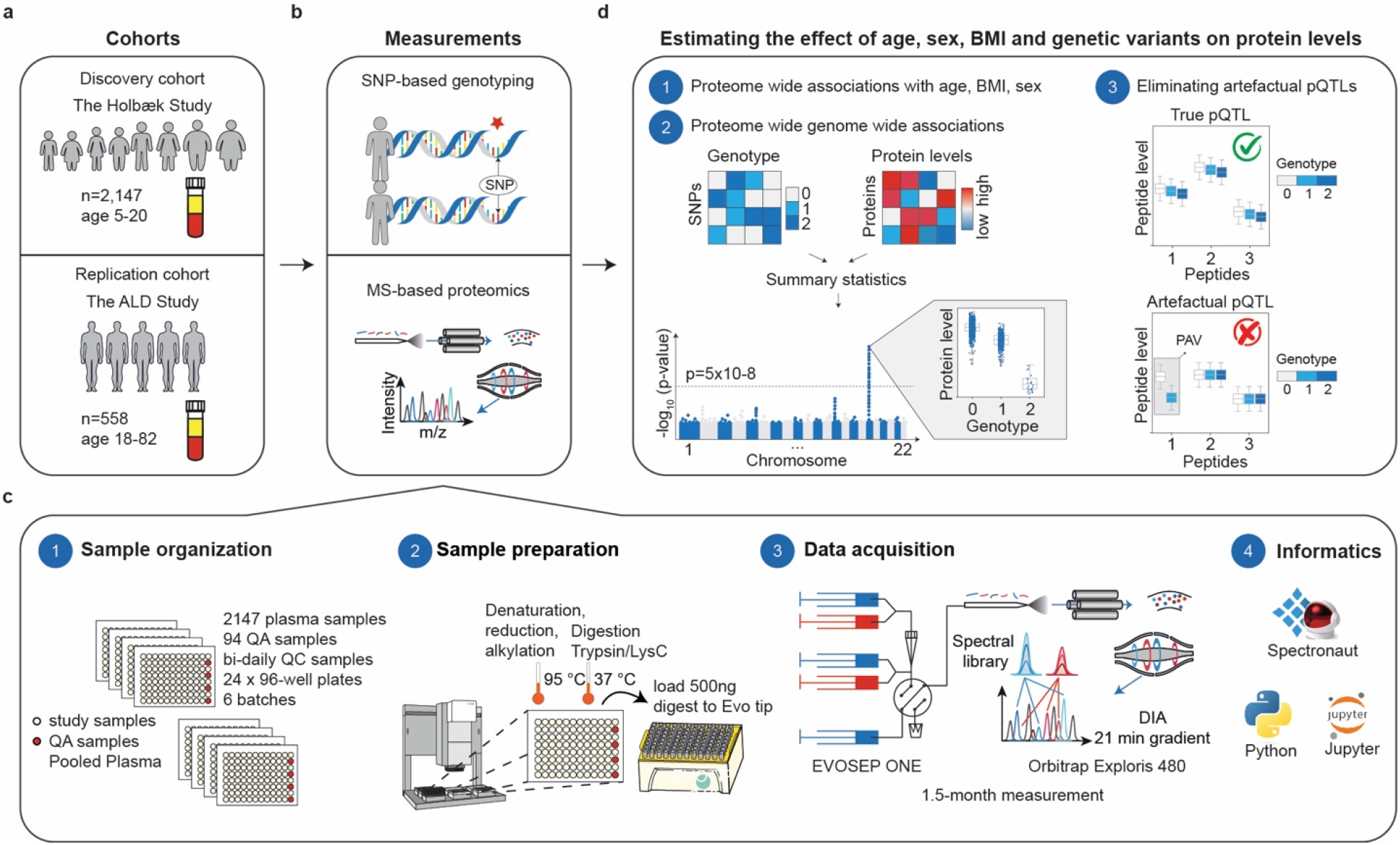
Study overview and proteomics workflow. **a**, Discovery- and replication cohorts used in this study. **b**, MS-based plasma proteome profiling and SNP-based genotyping were performed on both cohorts. **c**, Proteome profiling workflow and the computational tools used for processing of proteomics data including (1) sample organization, (2) sample preparation, (3) data acquisition and (4) informatics. **d**, Schematic representation of the analyses of associations between protein levels and age, sex, BMI SDS (1) and, genome-wide genetic variants (2) with a quality control step to eliminate artefactual pQTLs, as described below in the main text (3).

**Extended Data Fig. 1.**
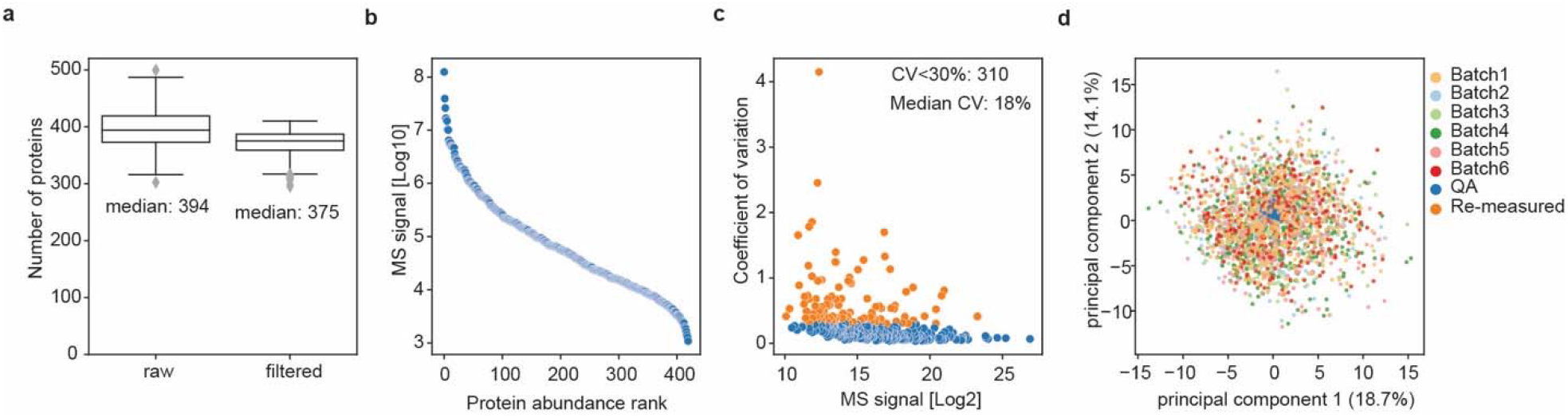
Proteomics data quality in the discovery cohort. **a**, The number of proteins quantified in each sample before and after filtering for data completeness at protein level. *n*=2,147 biologically independent samples. The gray line in the middle of the box is the median, the top and bottom of the box represent the upper and lower quartile values of the data and the whiskers represent the upper and lower limits for consideration of outliers (Q3□+□1.5□×□IQR, Q1□–□1.5□×□IQR). IQR represents the interquartile range (Q3□–□Q1). Median values are indicated. **b**, Protein intensity as a function of abundance rank. **c**, The coefficients of variation (CV) of each protein assessed by quality assessment samples are plotted against their median intensity. *n*=94 workflow replicates. **d**, Principal component analysis for the plasma proteome profile of all samples, showing the first and second principal component. Samples are color-coded according to the sample preparation batch.

## Nongenetic effects on the plasma proteome

To assess the impact of age, sex, and BMI SDS^22^ on the plasma proteome, we performed multiple linear regression analysis (Fig. 2a and Methods). As per the study design, we estimated the effect of BMI SDS separately for the group with overweight (BMI SDS >= 1.28) and the group with normal weight. Remarkably, 90% of quantified plasma proteins were associated with at least one of the three factors (about 60% with age, 43% with sex, and 63% with BMI SDS) and nearly a quarter of the quantified plasma proteome was associated with all three factors (Fig. 2b). Sex had the strongest associations in terms of effect size, followed by BMI SDS and age (Fig. 2c). Exploring the proteins most strongly associated with the three factors revealed those with known age-dependent effects such as F9^23^, RBP4^24^, and COL1A1^2^, and others not previously reported to be associated with age, such as GPLD1, APCS, and IGFALS (Fig. 2d). The association between IGFALS and age is supported by the fact that defects or low expression of IGFALS can lead to pubertal delay in children^25^. Similarly, we recapitulated known differences in protein levels between girls and boys, such as the pregnancy zone protein (PZP) and plasma esterase (BCHE), but also uncovered previously unreported proteins that exhibit sex-specific differences, including CD5L (Fig. 2f).

**Fig. 2.**
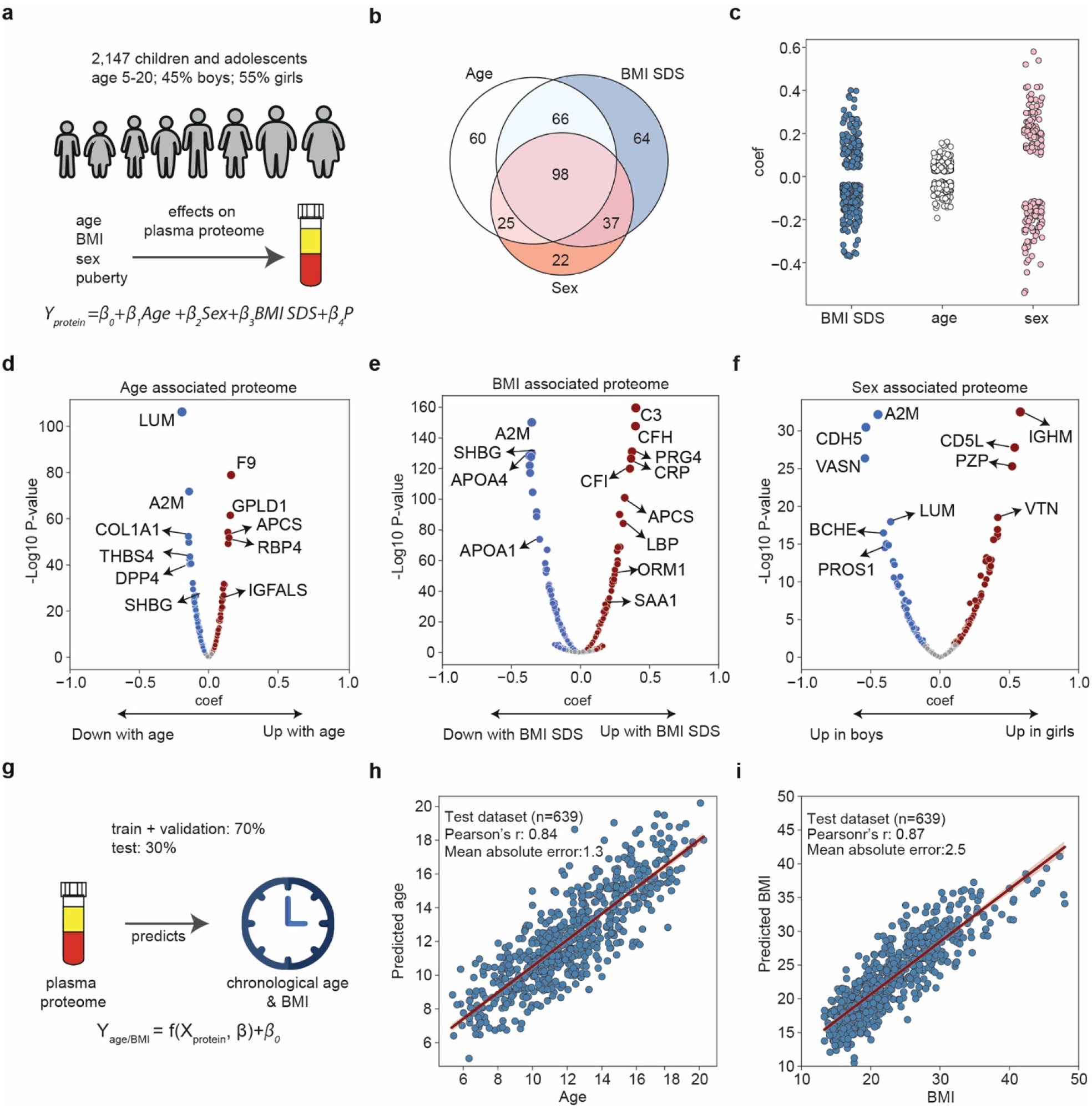
Relationship between the plasma proteome and age, BMI SDS, and sex. **a**, Schematic representation of linear modeling of the plasma proteome adjusted for pubertal stage and time to analysis. **b**, Number of proteins associated with age, BMI SDS and sex (FDR-corrected *p* < 0.05). *n*=1,603 biologically independent samples. **c**, Beta coefficients of proteins associated with age, BMI SDS, and sex. **d-f**, Volcano plots showing proteins associated with age (d), BMI SDS (e) and sex (f), highlighting strongly associated proteins. **g**, Schematic representation of linear modeling of age and BMI using plasma proteome. **h**,**i**, Prediction of age (h) and BMI in the test set (i). Pearson’s r between predicted and real values are indicated. *n*=639 biologically independent samples.

As expected, inflammatory proteins are most strongly associated with BMI SDS including the complement system proteins (C3, CFH, CFI), inflammatory markers (CRP), and acute phase proteins (APCS, SAA1, ORM1, LBP)^26^ (Fig. 2e). Interestingly, except for CFH and ORM1, all these proteins were statistically significant in the normal weight group as well, albeit with a smaller effect size, demonstrating that elevated levels of inflammatory proteins with increasing BMI SDS is not exclusive to obesity (Supplementary Table 2). Previously we had found PRG4 to decrease in response to weight reduction^26^, concordant with its positive association with BMI SDS in this data. PRG4 deficiency protects against glucose intolerance and fatty liver disease in mice, suggesting therapeutic potential of proteins identified here^27^.

Levels of plasma proteins can serve as a “biological clock” based on data from adults^2^. Having established the association between age and the plasma proteome in this cohort, we asked if the same is true in children and adolescents. Indeed, a set of 80 proteins predicted age with high accuracy (+/- 1.3 years, Pearson’s r 0.84 between predicted and actual age in a held out set of 639 individuals) (Supplementary Table 3, Fig. 2g-h and Methods). Likewise, a panel of 69 proteins accurately predicted BMI (Fig. 2i). Thus age, sex, and BMI SDS significantly contribute to the inter-individual variability in plasma protein levels, underscoring the impact of considering them in clinical studies of childhood diseases.

The few studies that have explored age-related trajectories in the plasma proteome, have primarily focused on early-stage development (infancy^4^ and childhood^3^) or adulthood^2^. Here we provide insights related to pre- and post-pubertal stages (age 5 to 20 years), addressing a gap in current knowledge. Hierarchical clustering analysis revealed seven age-related protein abundance trajectories, comprising between 5 and 38 proteins each (Fig. 3a, Supplementary Table 4 and Methods). Half of these proteins decrease with age. Notable examples include A2M in Cluster 1 with a linear trajectory, whereas THBS4 in Cluster 2 behaves non-linearly, remaining relatively constant until approximately age 12 years before declining eight-fold (Fig. 3c-e).

**Fig. 3.**
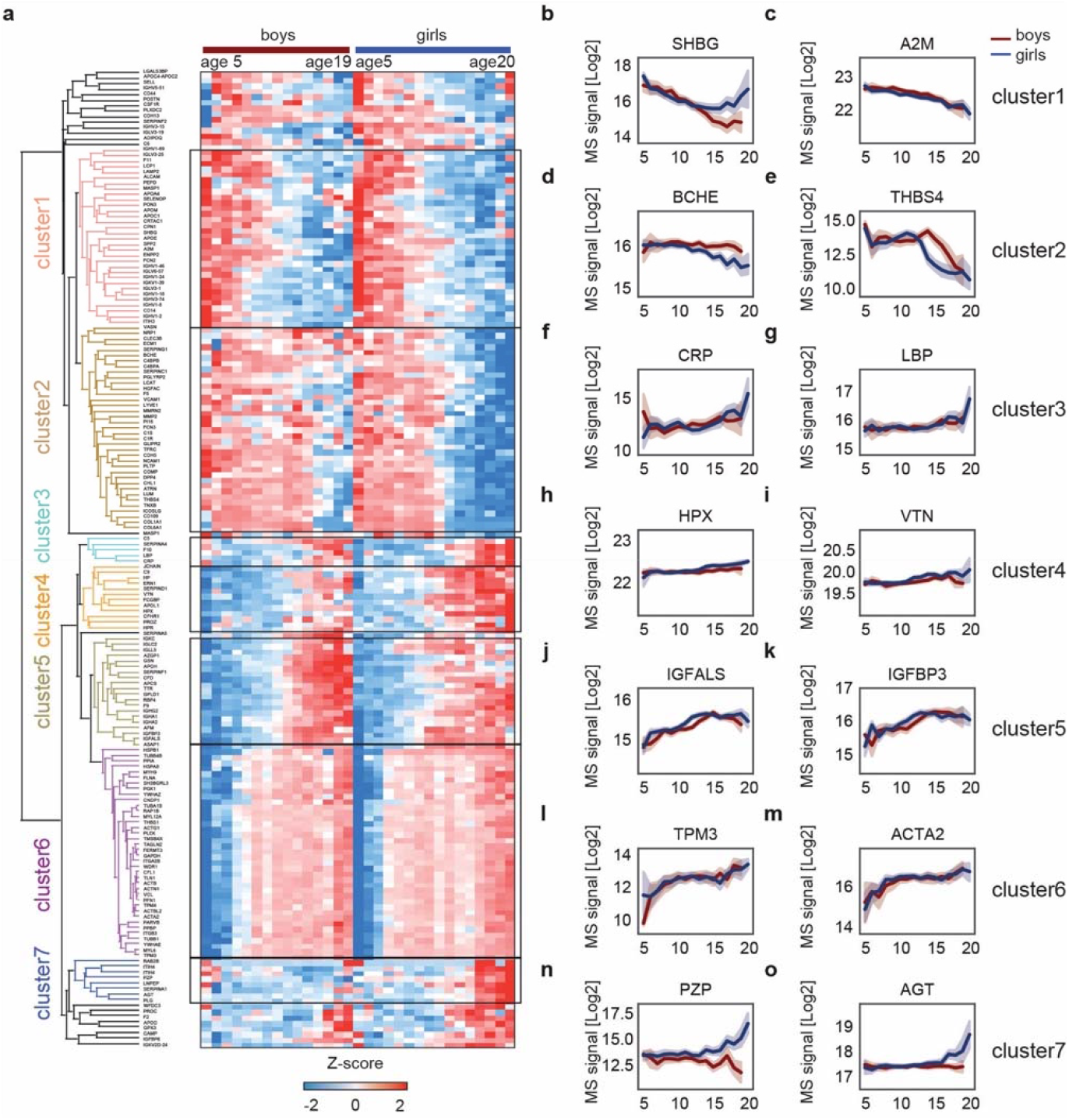
Sex-dependent temporal plasma proteome profiles. **a**, Hierarchical clustering dendrogram of proteins significantly associated with age. The heat maps display z-scored median intensities across age for girls (n=1,171) and boys (n=959). **b-o**, Temporal trajectories of representative proteins in Cluster 1-7 in panel (a). Mean values along the age axis and 95% confidence intervals are shown.

The overall trend of proteins in Clusters 3 and 4 is positive with age, such as LBP, HPX, and VTN (Fig. 2f-I). The well-established inflammatory marker CRP remained relatively stable until age 13 years before rising, which aligns with previous efforts to determine reference ranges of clinical parameters in children and adolescent populations^28^. Interestingly, the patterns of some proteins diverge between girls and boys at the onset of puberty, such as PZP and angiotensinogen (AGT) in Cluster 7 (Fig. 3n-o), as well as sex-hormone binding globulin (SHBG), the latter of which has been reported before^29^ (Fig. 3b). Girls reach puberty on average one year earlier than boys^30^, which is reflected by a corresponding shift in the trajectory of proteins such as IGFALS and IGFBP-3 (Fig. 3j-k).

A number of platelet proteins increased with age (Fig. 3i-m), which might have been caused by platelet contamination^31^. However, some of these proteins, including THBS1 and TLN1, have already been reported to have this trend^32^ and participants were randomly recruited regardless of age, making this observation unlikely a systematic bias. Taken together, our protein abundance trajectories may provide a valuable reference for investigating childhood diseases and establishing reference levels for development- and disease-relevant markers.

## Genetic effects on the plasma proteome

Next, we tested 5.2 million SNPs for association with plasma levels of 420 proteins in 1,914 individuals. We defined a primary pQTL as the most significant variant in linkage disequilibrium (LD) (r^2^>0.2) within a region (+/- 1Mb) that was associated with plasma levels of a protein (Methods). Applying a study-wide significance level of *p*<1.2×10^−10^ (5×10^−8^/420 proteins) resulted in 712 primary pQTLs for 158 proteins (Supplementary Table 5). For further downstream analysis, we decided to adopt the conventional GWAS significance threshold of *p*<5×10^−8^, resulting in 1,116 primary pQTLs and 201 regulated proteins (Fig. 4a). Genomic inflation was well controlled with median lambda_GC_=1.003 (standard deviation=0.004) (Methods). The identified pQTLs were distributed across chromosomes roughly as expected by their number of protein coding genes (Fig. 4b). These pQTLs are primarily located in non-coding regions, with only 3% and 1% being missense variants and synonymous variants, respectively (Methods, Fig. 4i and Supplementary Table 5). Remarkably, two thirds were in cis and more than 70% of the proteins had at least one cis-pQTL associated, implying pervasive local regulation (Fig. 4c-e). The MS-based proteomics data recapitulated that the same genomic locus can regulate multiple proteins, and that one protein can be regulated by multiple genomic loci. Specifically, 9% of the pQTLs were associated with more than one protein, while 72% of the proteins had multiple pQTLs associated with them – and 37% of these had pQTLs located on different chromosomes (Fig. 4f-g).

**Fig. 4.**
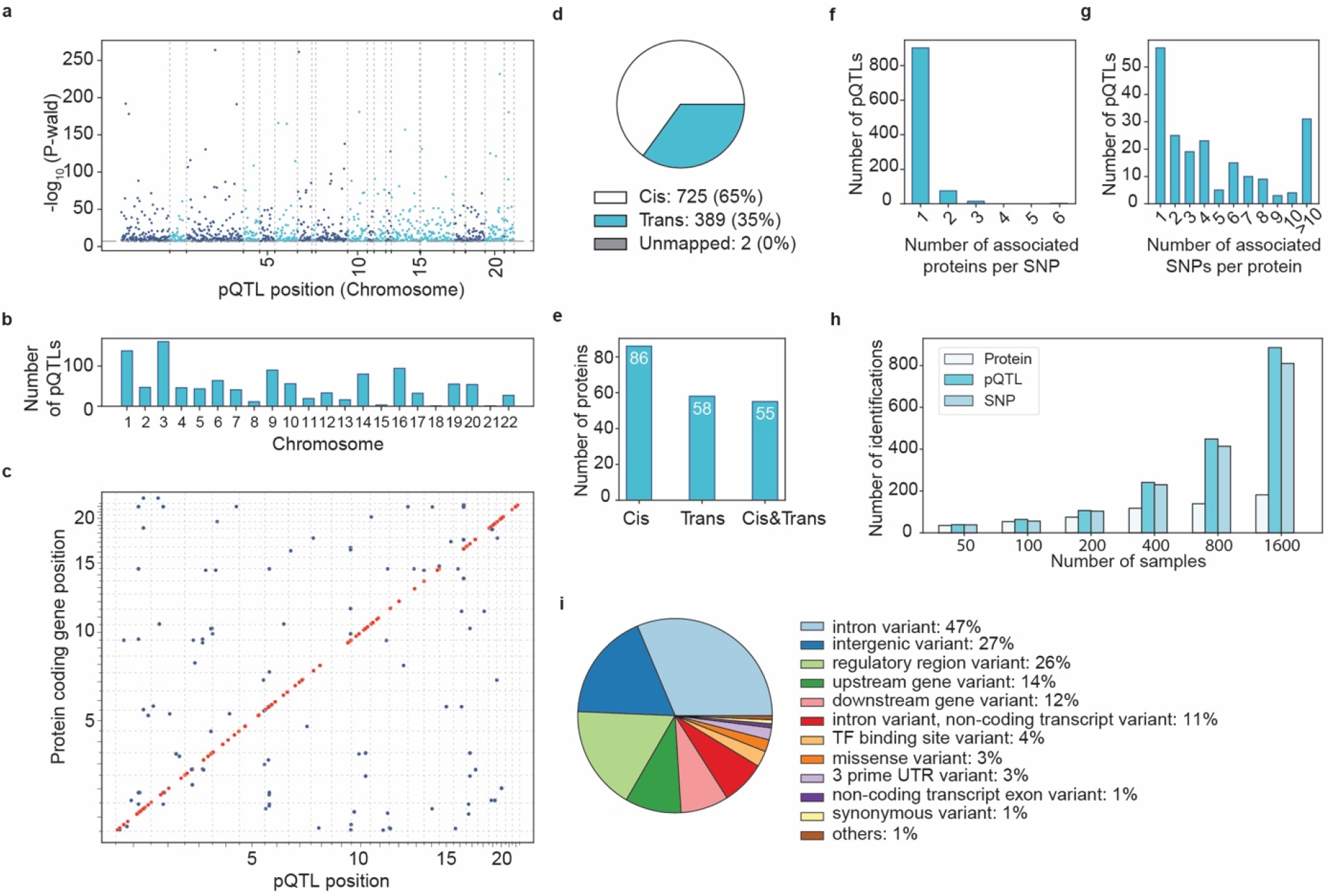
Genetic architecture of pQTLs. **a**, Primary pQTLs across the genome. **b**, The number of primary pQTLs by chromosome. **c**, Primary pQTLs against the locations of the transcription start site of the gene coding the protein target. Cis- (red) and trans- (blue) pQTLs. **d**, The number of cis- and trans pQTLs. **e**, The number of proteins that are associated with cis only-, trans only- and both cis and trans pQTLs. **f**, Distribution of number of associated proteins per SNP. **g**, Distribution of number of associated SNPs per protein. **h**, Identification rate of pQTLs with increasing sample size. **i**, Variant annotation.

Compared to previous MS-based pQTL studies in blood, at study-wide significance level (*p*<1.2×10^−10^), our study identified pQTLs for 2.5 to 30 times more proteins^14–17^. Simulation suggested that this was largely due to limited sample sizes (Fig. 4h). In comparison to affinity-based studies of similar size, we also identified pQTLs for a higher fraction of proteins analyzed^33–35^. A greater efficiency at identifying pQTLs, could reflect the ultra-high specificity of identification that is a hallmark of MS-based proteomics. However, our sample sizes are up to 25-fold less than those of the latest affinity-based projects^8–10^. Thus, a next step should be to perform pQTL studies on tens of thousands of samples and also with increasing depth, perhaps with enrichment strategies^36^. Nonetheless, our study already identified pQTLs for 50% of the analyzed proteome, a proportion that may even go up with sample size, revealing extensive genetic influence over the plasma proteome.

## Eliminating artefactual pQTLs

Protein variants can modify binding surfaces and lead to alterations in peptides encompassing variant regions, potentially introducing biases in proteomic studies^11,17,37^. We developed a framework to thoroughly examine peptide-level information for indications of artefactual pQTLs (Extended Data Fig. 2a). Taking into account that not all peptides of a protein are detected, and that algorithms will not use all peptides for quantification, we flag a pQTL as a false positive if the signal arose solely from the peptide that is affected by protein-altering variants with no corroborating evidence from other peptides (Methods). Among all plasma proteins with pQTLs, only one protein presented artefactual pQTLs. Specifically, the measurement for the Complement Factor H (CFH) protein (Uniprot ID A0A0D9SG88) was biased due to the missense variant rs1061170, which results in the substitution of histidine with tyrosine (H/Y) at position 402. This amino acid change introduces an artificial difference in protein levels across the genotypes (Extended Data Fig. 2b-c). Although in rare cases, missense variants could influence the accuracy of the effect size measured, they did not result in false identifications of pQTLs (Extended Data Fig. 2d-e). Consequently, more than 98% of the pQTLs were reliable, demonstrating a high level of confidence in the results obtained through mass spectrometry (MS)-based proteomics.

**Extended Data Fig. 2.**
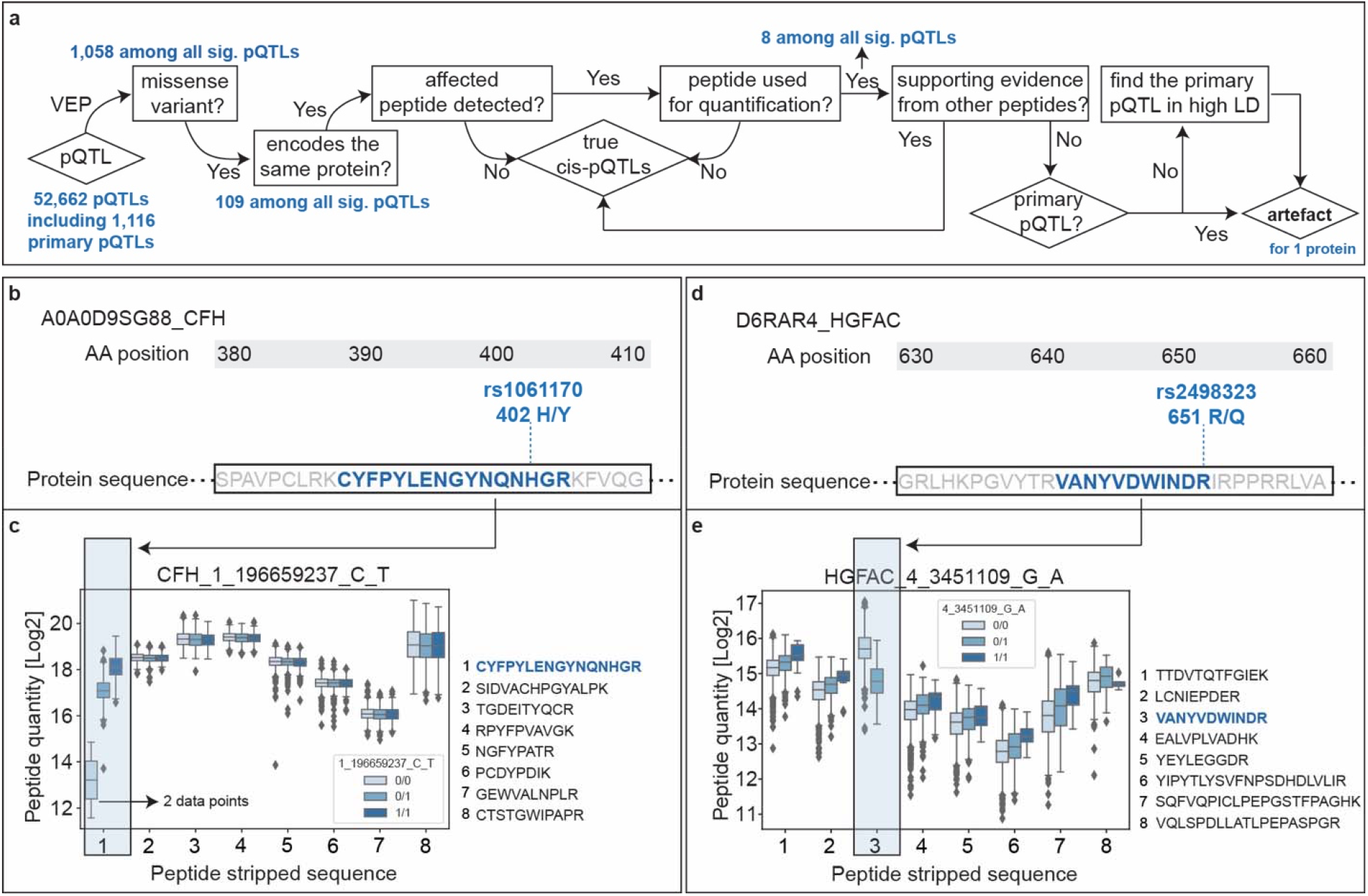
Peptide-level data distinguishes true pQTLs from artefacts. **a**, A framework to identify artefact pQTLs caused by missense variants. **b**, Protein sequence of CFH (Uniprot ID A0A0D9SG88) that contains the amino acid affected by the missense variant rs1061170. **c**, Peptide level quantification of CFH in individuals stratified by rs1061170. **d**, Protein sequence of HGFAC (Uniprot ID D6RAR4) that contains the amino acid affected by the missense variant rs2498323. **e**, Peptide level quantification of HGFAC in individuals stratified by rs2498323.

## Characterization of pQTL effect sizes

Having established the quality of the pQTLs, we investigated the effect size based on beta statistics derived from the association tests as well as allelic fold change calculated on data without normalization^38^ (Methods). While they were mostly mild, 79 pQTLs for 27 proteins were associated with two- to 35-fold change in protein levels comparing homozygous genotypes for the reference (0/0) and alternative (1/1) allele (Fig. 5a-b and Supplementary Table 6). Conversely, relative changes in protein levels of 6% corresponding to the smallest absolute beta value were also confidently detected. Local (cis-pQTL) regulation generally had the largest effect sizes (Extended Data Fig. 3a-b). Furthermore, variants located in the 5’ and 3’ untranslated regions, non-coding exons as well as transcription factor binding sites had a larger average effect size compared to those located in the intron and intergenic regions (Extended Data Fig. 3c), supporting an important role of these regions in transcriptional regulation. Large genetic effect sizes for proteins such as HLA-C, MST1, C4A, PROCR, LBP, and CFHR may have important implications for clinical and biomarker research (Fig. 5c-h). For instance, we found a protein coding variant as a pQTL that reduced levels of lipopolysaccharide (LPS)-binding protein (LBP) four-fold (Fig. 5g). As this protein is important in innate immunity, we speculate that individuals with the mutant form have compromised immunity, which has indeed been reported^39^. The observed substantial genetic effects emphasize the importance of considering pQTL information when interpreting findings from clinical and biomarker research, particularly for proteins whose plasma levels are under strong genetic control.

**Fig. 5.**
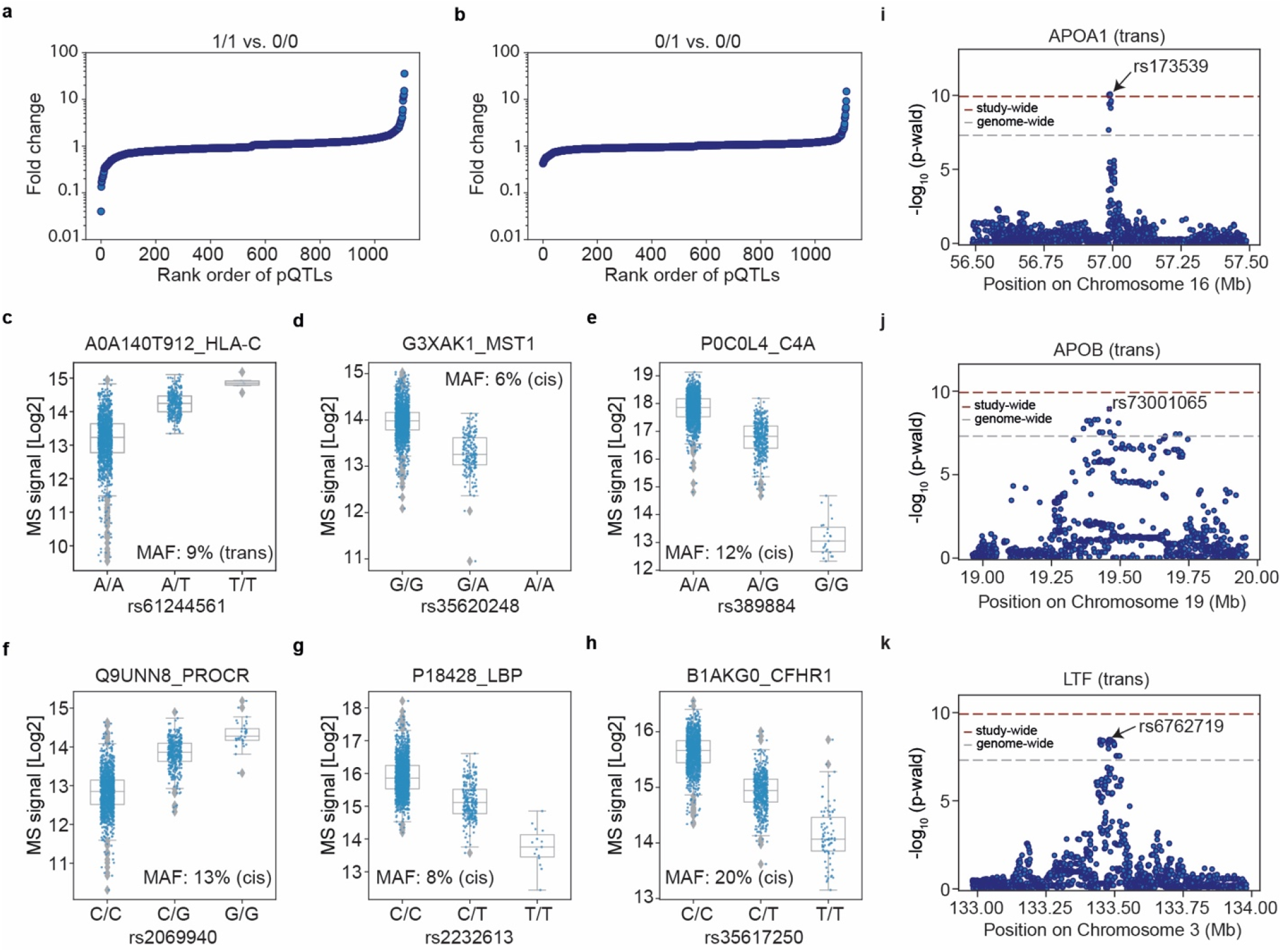
Effect sizes and integration of pQTLs with known variant-trait associations. **a**, Fold change of protein levels between homozygous variant alleles and homozygous reference alleles for all primary pQTLs. **b**, Fold change of protein levels between heterozygous and homozygous reference alleles for all primary pQTLs. **c**,**d**,**e**,**f**,**g**,**h**, Distribution of log2 intensity values of the top six proteins with the highest absolute beta value. The gray line in the middle of the box is the median, the top and bottom of the box represent the upper and lower quartile values of the data and the whiskers represent the upper and lower limits for consideration of outliers (Q3□+□1.5□×□IQR, Q1□–□1.5□×□IQR). IQR represents interquartile range (Q3□–□Q1). For genotype 0/0:0/1:1/1, *n*=1278:328:6, 1708:191:0, 1490:399:23, 1225:410:35, 1637:260:15, 1227:606:79, respectively. Only non-imputed values are shown. **i**,**j**,**k**, Regional association plots between APOA1 and trans locus rs173539 (i), APOB and trans locus rs73001065 (j), and LTF and trans locus rs6762719.

**Extended Data Fig. 3.**
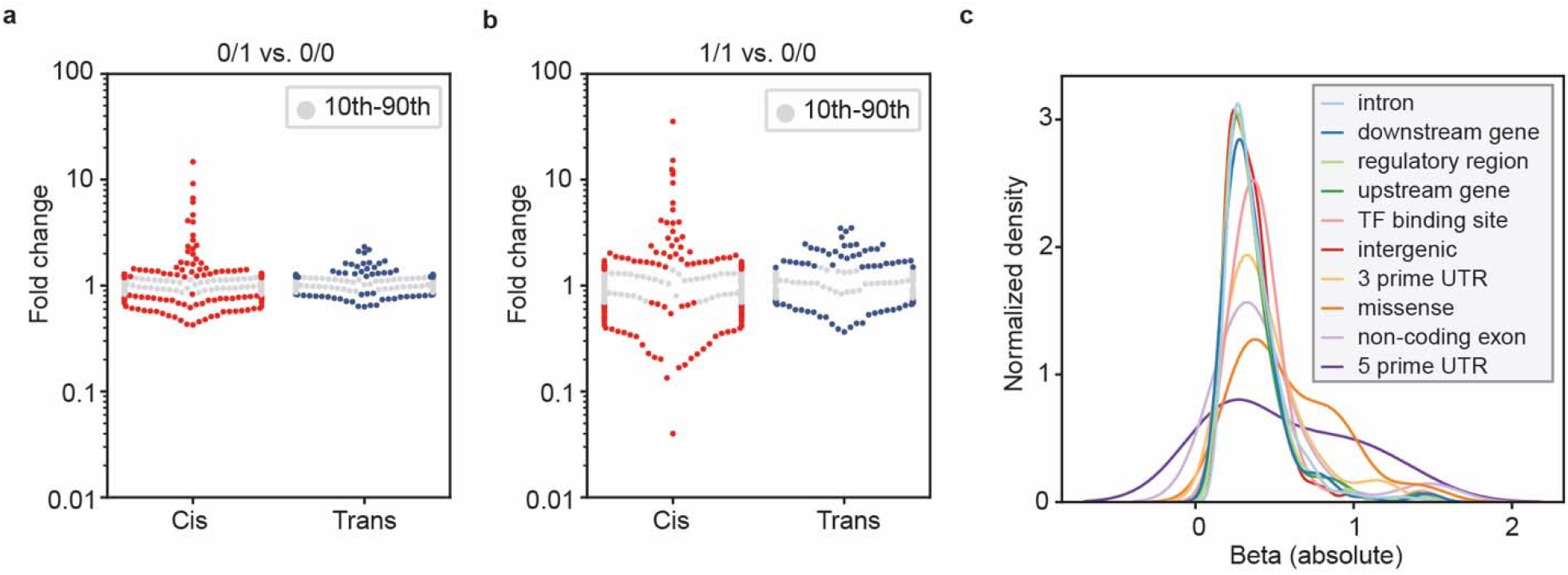
Effect size according to pQTL categories. **a**, Swarmplot showing fold change of protein levels between homozygous variant alleles and homozygous reference alleles for all primary pQTLs stratified by cis- and trans-pQTLs. Values within 10^th^-90^th^ percentiles are indicated. **b**, Fold change of protein levels between heterozygous variant alleles and homozygous reference alleles for all primary pQTLs stratified by cis- and trans-pQTLs. **c**, Kernel density estimation plot of beta statistics stratified by variant categories.

## Novel pQTLs link proteins to GWAS results

To investigate if our MS-based study had discovered any novel pQTLs, we compared it to 33 published studies (Extended Data Fig. 4a, Supplementary Table 7 and Methods). This revealed 310 novel pQTLs regulating 93 proteins, 47 of which had no previously reported genetic regulation. This includes, for instance, the C4A and C4B isotypes of complement factor 4, which cannot be distinguished by affinity-based platforms (Supplementary Table 5). Their sequences differ in less than one percent, but this greatly affects their binding affinity towards their molecular targets^40^. Of the remaining pQTLs, 61% were replicated in at least five studies (Extended Data Fig. 4b).

Genome wide association studies (GWAS) provide increasingly detailed associations between genomic loci and phenotypes but are missing a mechanistic link to the proteins mediating the effect^38,41,42^. With a view to close this gap, we next investigated whether any of our novel pQTLs were known to be associated with phenotypic traits. Mapping our protein-associated variants to the GWAS Catalog revealed 85 such cases (Supplementary Table 8 and Methods). Many of these were immediately biologically plausible, for example the trait ‘high density lipoprotein (HDL) cholesterol levels’ is associated with rs173539, a variant that our data connects to levels of APOA1, a component of HDL (Fig. 5i). This suggests that the variant influences HDL cholesterol levels by upregulating APOA1 levels. Similarly, rs73001065 has been associated with a decreased risk for non-alcoholic fatty liver disease, which our data indicates may happen through reduced levels of APOB (Fig. 5k). Finally, the ion binding capacity associated variant rs6762719 regulates levels of the transferrin protein LTF (Fig. 5j).

Cross-referencing all significant pQTLs to GWAS results, beyond the novel ones, provides many more links between variant and phenotypic trait associations (Supplementary Table 8). Some but not all of these connections are expected, but others open up intriguing avenues for further investigation. These results illustrate how integrating high-confidence pQTLs with GWAS results helps to uncover or understand the molecular mechanisms between variant-disease/trait associations.

**Extended Data Fig. 4.**
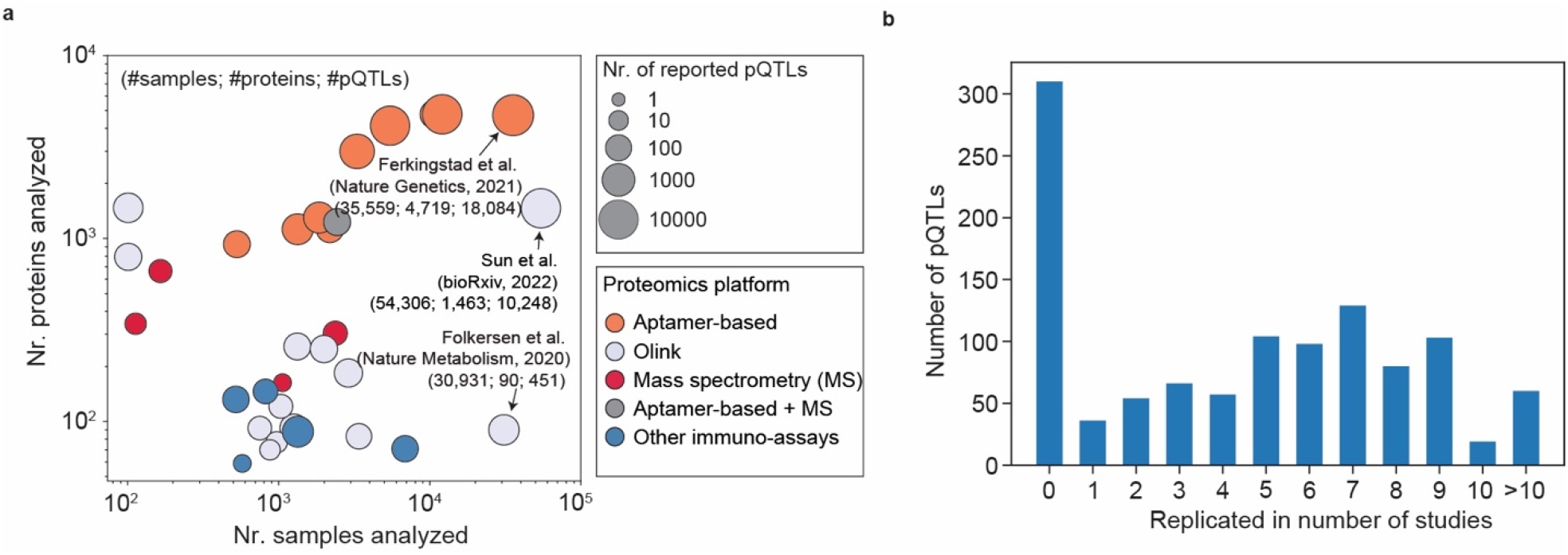
Comparison of pQTLs to previous plasma or serum studies. **A**, Comparison of the number of proteins analyzed and the number of samples analyzed by 33 previous studies. Studies were color-coded based on the proteomics platforms used to generate the proteomics data. **B**, The number of pQTLs replicated in previous studies.

## Highly replicated pQTLs in an independent cohort

To test if our primary pQTLs could be replicated in an independent cohort with a completely different age range and other phenotypic characteristics, we leveraged an existing adult cohort of alcohol-related liver disease (ALD) that we had recently measured by MS-based proteomics^18^. Of the 93% of pQTLs that could be compared due to genotype and proteomics quality filters and at a nominal significance level of *p*<0.05, a full 90% were successfully replicated – 94% of the cis, 83% of the trans, and 72% of the novel (Fig. 6a, Supplementary Table 9, Methods). Furthermore, the direction and magnitude of the effects aligned well between discovery and replication cohorts (Pearson’s r=0.97, Fig. 6b), which is also reflected by the strikingly similar distribution of protein levels among genotypes between the two cohorts – see Extended Data Fig. 5a-I for examples.

**Fig. 6.**
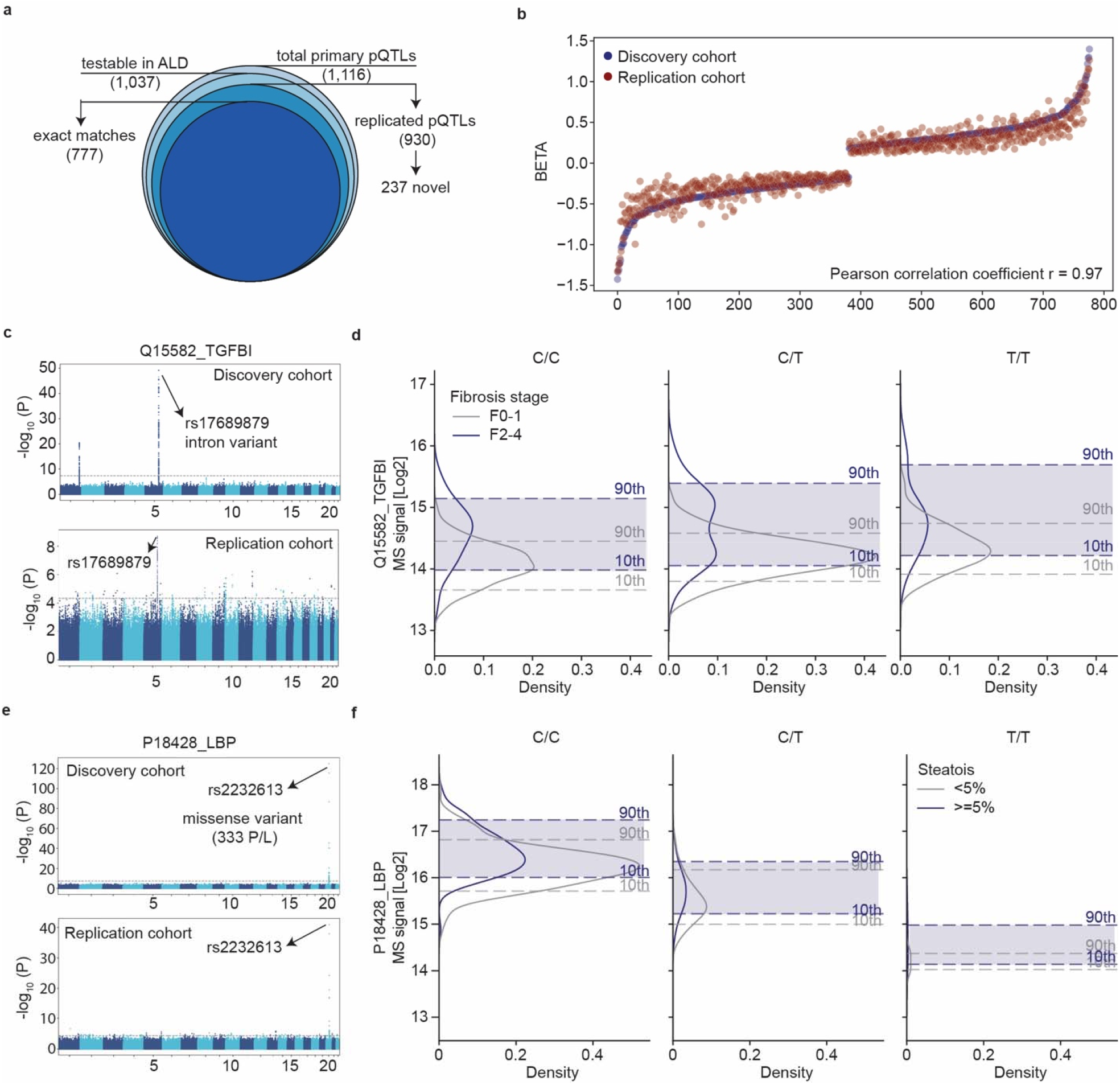
Replication of pQTLs in an independent cohort. **a**, Testable and replicated pQTLs in the independent study. **b**, Beta values of pQTLs in the discovery and replication study, with pQTLs ranked in increasing order of beta values in the discovery study. Pearson’s r of beta values between the two cohorts is indicated. **c**, Manhattan plot of association between SNPs and plasma levels of TGFBI in the discovery cohort (upper panel) and replication cohort (lower panel) with the lead variant annotated. **d**, Distribution of plasma levels of TGFBI stratified by the genotype of its pQTL and fibrosis stage in the replication cohort. For genotype 0/0:0/1:1/1, n=96:181:90 and 55:93:43 for fibrosis stage f0-1 and F2-4, respectively. **e**, Manhattan plot of association between SNPs and plasma levels of LBP in the discovery cohort (upper panel) and replication cohort (lower panel) with the lead variant annotated. **f**, Distribution of plasma levels of LBP stratified by the genotype of its pQTL and steatosis stage in the replication cohort. For genotype 0/0:0/1:1/1, n=318:52:3 and 157:25:3 for steatosis <5% and ≥5%, respectively.

We next asked if pQTL information could improve biomarker performance. We had previously reported three marker panels for detecting liver fibrosis, inflammation and steatosis ^18^. For half of these, including TGFBI and LBP, which had the largest genetic effect sizes, our study identified replicated pQTLs (Fig. 6c and Fig. 6e). Our data revealed a shift in distribution of TGFBI protein levels depending on its corresponding pQTL in both disease and control groups (Fig. 6d). Adding genotype information to protein levels further improved the accuracy of classifying patients with F0-1 vs. F2-4 fibrosis stages using TGFBI (Extended Data Fig. 6a). Similarly, we observed a more striking downward shift in LBP levels in the presence of the variant rs2232613 (Fig. 6f, Extended Data Fig. 6b). These findings suggest that pQTLs should be considered in biomarker research, especially for those with strong genetic effects. However, whether integrating pQTLs in classification algorithms has beneficial effects needs to be evaluated on a case-by-case basis.

**Extended Data Fig. 5.**
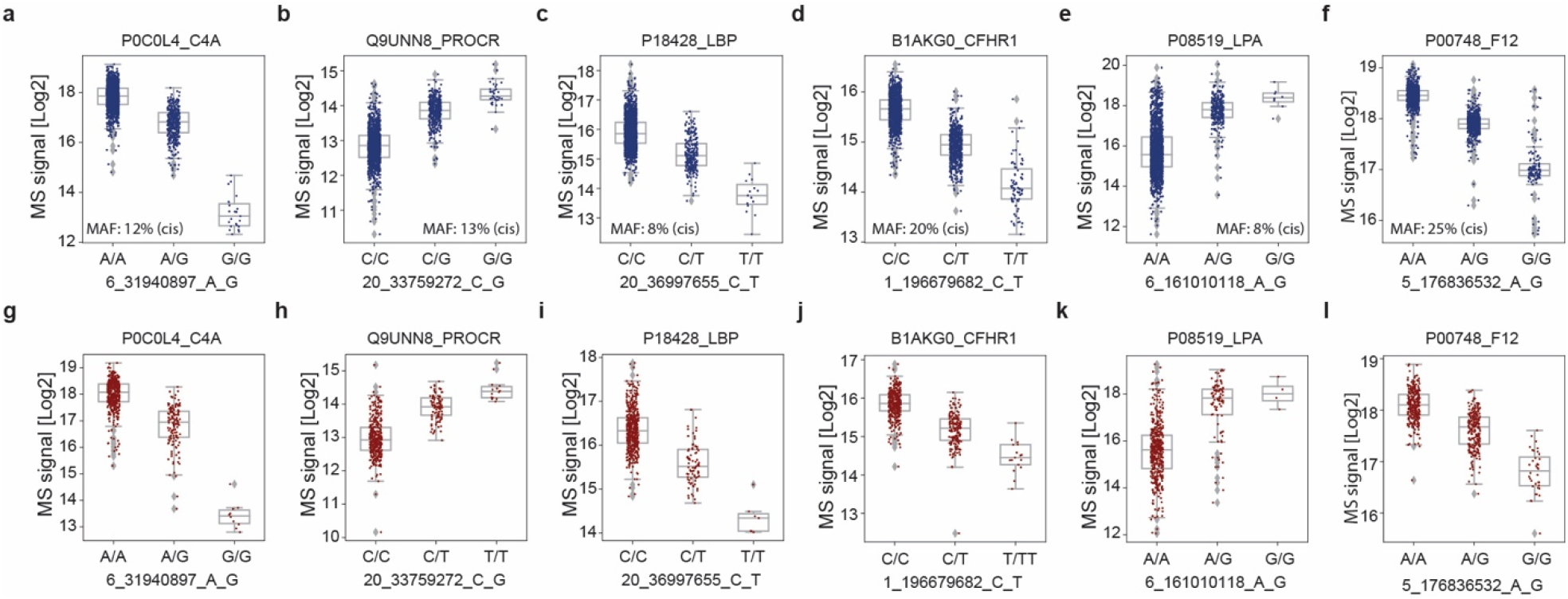
Replication of pQTLs in an independent cohort. **a-l**, Distribution of log2 intensity values of the top six proteins with the highest absolute beta value in the discovery study (a-f) and replication study (g-i). The gray line in the middle of the box is the median, the top and bottom of the box represent the upper and lower quartile values of the data and the whiskers represent the upper and lower limits for consideration of outliers (Q3□+□1.5□×□IQR, Q1□–□1.5□×□IQR). IQR represents interquartile range (Q3□–□Q1). For genotype 0/0:0/1:1/1, *n*=1490:399:23, 1225:410:35, 1637:260:15, 1227:606:79, 1602:288:8, 1078:716:118 for a-f, and *n*=427:133:10, 373:105:14, 482:82:7, 374:179:17, 458:91:4, 303:232:36 for g-l, respectively. Only non-imputed protein values are shown.

**Extended Data Fig. 6.**
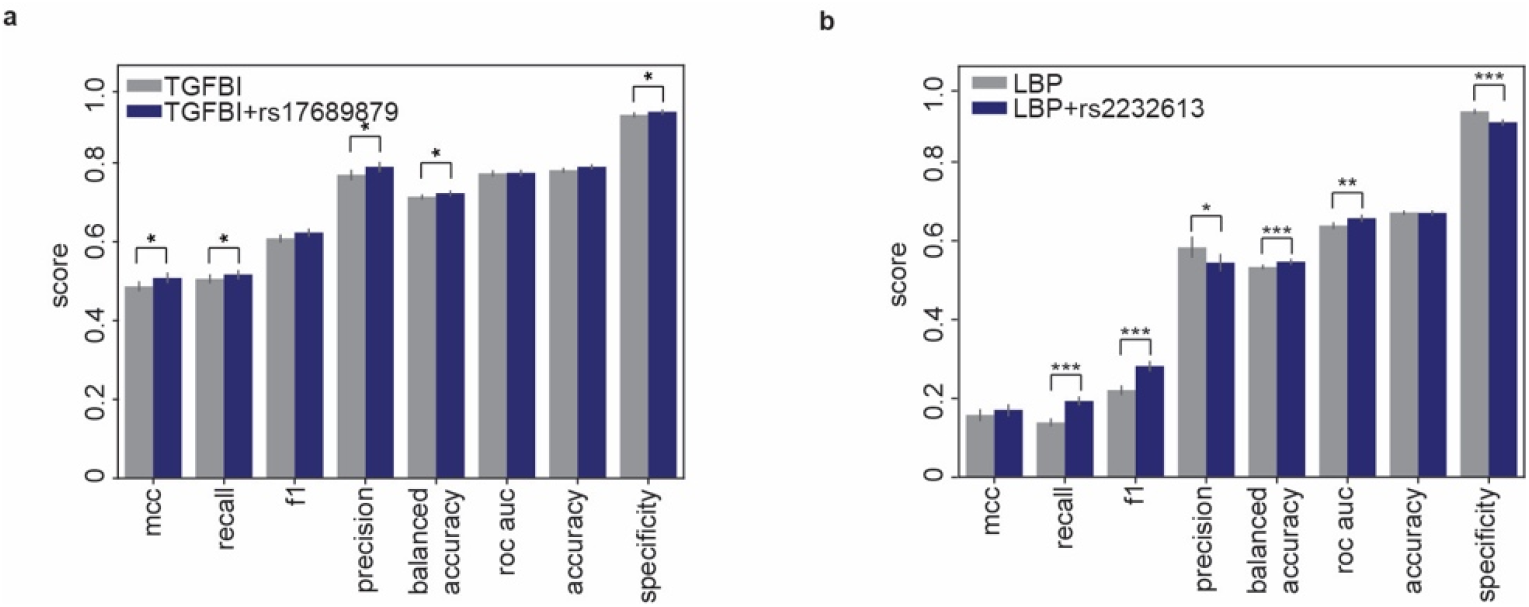
Incorporating pQTLs affect biomarker performance. **a**, Classification performance metrics of TGFBI and TGFBI+rs17689879 for identifying significant fibrosis in the ALD cohort^18^. Error bars represent s.d. Significance levels are indicated (* *p*<0.05, ** *p*<0.01, *** *p*<0.001) **b**, Classification performance metrics of LBP and LBP+rs2232613 at identifying any steatosis.

## Discussion

Here, in one of the most extensive MS-based proteomics studies to date, we analyzed the plasma proteomes of more than 2,000 children and adolescents, obtaining highly specific and quantitative protein trajectories throughout childhood and adolescence. Some of these age-dependent trajectories exhibit sex-specific patterns reflective of puberty-related differences between girls and boys. One striking finding was that about 90% of all quantified plasma proteins were significantly regulated – about half of these by genetic variants. Turning these relationships around allowed accurate prediction of the age or BMI from the plasma proteome. The results of our study can be explored online and are a resource for understanding protein changes across age and sex, and can serve as a reference for proteomic biomarker studies of childhood diseases (proteomevariation.org).

Our study emphasizes the importance of large sample sizes in pQTL studies, which has already been made for non-MS-based proteomics^8^. We demonstrated by simulation that previous MS-based studies would not have been expected to yield many pQTLs for statistical reasons. Although the largest of its kind, its size is still moderate compared to affinity-based studies. Nevertheless, we found evidence for regulation by genetic variants for at least half the quantified plasma proteome, as well as hundreds of novel pQTLs.

The high replication rate in the independent adult cohort and the highly consistent direction and strength of effects between the two cohorts suggest that pQTLs between children and adults are comparable, despite plasma protein levels being influenced by many non-genetic factors, such as disease. Some effect sizes were particularly large – often several-fold – suggesting important physiological consequences. Furthermore, our results indicate a need to tailor the reference levels of candidate biomarkers according to pQTLs affecting the protein of interest. We found that incorporating pQTLs of candidate biomarkers can improve their classification performance. This may apply to prognostic and predictive biomarkers as well.

Our study also has limitations. First, it employs a cross-sectional design. A longitudinal study design would reduce inter-individual variability between time points and thus more accurately depict the age-dependent trajectories of protein levels in plasma. Second, we performed stringent quality control on the genotype data, and nearly 90% of total SNPs were filtered out due to low minor allele frequency and/or imputation quality. Consequently, low frequency pQTLs may have been omitted. A larger sample size is needed to lower the minor allele frequency threshold. Third, we could not exclude the possibility that unobserved SNPs could be protein-altering variants in high linkage disequilibrium with the identified pQTLs and therefore contribute to artefactual pQTLs.

Existing large-scale pQTL studies have predominantly used affinity-based proteomics platforms, which are optimized for body fluids. These platforms report the quantification of tens to thousands of proteins, but they do not always agree and large-scale validation of the specificity of binding reagents is lacking, ideally obtained by orthogonal methods such as MS^11–13^. Additionally, although artefactual pQTLs due to the ‘epitope effect’ associated with these platforms can be estimated by approaches such as conditional analysis and comparing pQTLs to expression QTLs, they cannot be directly assessed^7,10^. In contrast, MS-based proteomics has the advantages of being highly specific and agnostic to the sample type and species, and we here show how artefactual pQTLs can be directly eliminated using peptide-level data. Our data also indicates that MS-based proteomics more efficiently retrieves pQTLs than affinity-based approaches at similar sample sizes. It would now be exciting to study this with further comparative analysis, accounting for various factors such as cohort characteristics, computational methods, sample size, proteome coverage, and variants analyzed. Clearly, there is a need to further broaden the proteome depth without compromising throughput and reproducibility, which could be accomplished by emerging workflows that combine depletion- and multiplex strategies as well as new MS acquisition schemes.

To date, affinity-based studies have identified tens of thousands of pQTLs for thousands of circulating proteins. However, the quest for pQTL discovery is far from complete especially regarding sample types beyond plasma. Many proteins are only leaked into the plasma, but have their primary role in tissues, where MS-based proteomics can readily quantify nearly complete proteomes. A few pioneering MS-based proteomics studies with a few hundred samples in human or mouse have already identified pQTLs in the human brain and liver proteomes^43–45^. Based on the results shown here, we propose to greatly expand such efforts. Besides establishing a deep and high-throughput workflow, access to genotyped samples will be a key challenge in this exciting endeavor. Beyond bulk tissue, specific cell types in specific tissues could also be envisioned^46^. We believe these efforts will enable fruitful downstream applications such as colocalization and Mendelian Randomization for causal inference and identification and prioritization of drug candidates^47,48^.

## Method

### Study participants

We included 2,147 children and adolescents (55% girls and 45% boys, 17 had missing values) from The HOLBAEK Study, aged 5 through 20. The participants were recruited from 1) the Children’s Obesity Clinic, Centre of Obesity Management offering the multidisciplinary childhood obesity management program at Copenhagen University Hospital Holbæk^49^ and 2) a population-based cohort recruited from schools in 11 municipalities across Zealand, Denmark^50^ in a cross-sectional study design. Both groups were enrolled between January 2009 and April 2019. Eligibility criteria for the children in the obesity clinic group were an age of 5–20 years and a BMI above the 90^th^ percentile (BMI SDS >=1.28) according to Danish reference values^22^. Exclusion criteria for this study for both groups are 1) age at recruitment younger than 5 years or older than 20 years; 2) diagnosed type 1 diabetes; 3) diagnosed type 2 diabetes; 4) treatment with medications including insulin, liraglutide, and/or metformin; 5) meeting type 2 diabetes criteria^51^ based on the blood sample taken for this study (fasting plasma glucose > 7.0 mmol/L and/or hemoglobin A1c (HbA1c) > 48 mmol/mol. The study protocol was approved by the ethics committee for the Region Zealand (protocol no. SJ-104) and is registered at the Danish Data Protection Agency (REG-043-2013). The HOLBAEK Study including the obesity clinic cohort and the population-based cohort are also registered at ClinicalTrials.gov (NCT00928473). The study was conducted according to the principles of the Declaration of Helsinki, and oral and written informed consent was obtained from all participants. An informed oral assent was given by the participant if the participant was younger than 18, and the parents gave informed written consent. Tanner stage^52,53^ was evaluated by a pediatrician for individuals recruited at the obesity clinic and self-evaluated using a questionnaire with picture pattern recognition for individuals in the population-based group.

### Plasma proteomics

We prepared 2,147 plasma samples together with 94 quality assessment samples (pooled plasma sample) on an automated liquid handling system (Agilent Bravo) in a 96-well plate format as previously described^18,54^ in six batches. We acquired plasma proteomics data using a DIA method and the Evosep One liquid chromatography system coupled online to an Orbitrap Exploris 480 mass spectrometer as previously described^18,54^ between August and September in 2021 (6-week measurement time in total). We analyzed the plasma proteomics dataset with Spectronaut v.15.4^55^. Default settings were used unless otherwise noted. Data filtering was set to ‘Qvalue’. ‘Cross run normalization’ was enabled with the strategy of ‘local normalization’ based on rows with ‘Qvalue complete’. FDR was set to 1% at both the protein and peptide precursor levels. A previously generated deep fractionated plasma data-dependent acquisition (DDA) library was used in the targeted analysis of DIA data against the human reference proteome database (2018 release, 21,007□canonical and 72,792□additional sequences). Plasma proteomics dataset was filtered for 40% valid values across all samples (proteins with >60% missing values were excluded from downstream statistical analysis), log2 transformed with the remaining missing values imputed by drawing random samples from a normal distribution with downshifted mean by 1.8 and scaled s.d. (0.3) relative to that of abundance distribution of all proteins in one sample. Specifically, in total 588□proteins were quantified, filtering for 40% valid values across all samples resulted in a dataset of 420□proteins with a data completeness of 88%. Assessed on 94□quality assessment samples, median workflow coefficient of variation was 18% across a 6-week measurement period (Extended Data Fig. 1). The resulting dataset was then corrected for sample preparation batches using ComBat (version 0.3.2)^56^.

### Association of plasma proteome with age, sex, and BMI SDS

Plasma protein levels were normalized by rank-based inverse normal transformation (INT) with a Python implementation (https://github.com/edm1/rank-based-INT). The default Blom offset of c=3/8 was adopted. We used the following equation:

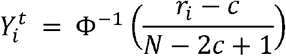

where *r*_*i*_ is the rank of the *i*th observation among total number of N, and Φ^−1^ denotes the quantile function (or percent point function) implemented in SciPy (version 1.7.1; https://scipy.org/citing-scipy/). We used multiple linear regression implemented in Pingouin^57^ (version 0.4.0) to estimate the effects of age, sex, BMI SDS^22^ on the protein level. We included pubertal status and time to analysis (plasma sample storage) as covariates in the regression model. Pubertal status was dichotomized according to the tanner stage: tanner 1 = prepubertal, tanner 2-5 =pubertal/post-pubertal. Associations were considered significant if the Benjamini-Hochberg corrected p-values were below 0.05. For 83% of the proteins, residuals are normally distributed (Shapiro-wilk test). Samples with missing values of variables in the regression analysis were excluded (n=1,603 remained).

### Prediction of age and BMI using plasma proteins

We used the linear regression model implemented in scikit-learn^58^ (sklearn version 1.0) for the prediction of age and BMI (here we used the BMI without adjusting for age and sex). We split the dataset into a training and test set (70:30) with the training set further divided into a training and validation set (70:30). A linear regression model was trained using all protein features in the training set. Features were ranked based on their coefficients in absolute values and a new model was trained again with an increasing number of features from one to 420 on the training set and evaluated on the validation set where the optimal number of features was determined based on the mean squared error. The final model was then trained on the training set using the selected features and evaluated on the held-out test set. Mean absolute error and Pearson’s r between the predicted values and real values in the test set were calculated to indicate prediction accuracy.

### Hierarchical cluster analysis of protein trajectories

Unsupervised hierarchical clustering of age-associated proteins was performed in the Perseus computational software (1.6.5.0). Proteins that passed the Benjamini-Hochberg corrected P-value with an absolute coefficient of above 0.04 were included. Row clustering was based on median log2-intensity after Z-score normalization across ages for girls and boys, respectively.

### Genotyping and imputation in the discovery cohort

Participants in this study were genotyped in three batches on the Infinium HumancoreExome12 v1.0 and HumancoreExome24 v1.1 Beadchips (Illumina, San Diego). Genotypes were called using the Genotype module of the GenomeStudio (Illumina). Before imputation, datasets from the 3 different batches were merged after quality control (only variants present on both chip versions were kept), and monomorphic variants as well as batch-associated variants were removed (Fishers exact test, P < 1e-7). We used the Sanger imputation server to phase the genotype data using EAGLE2 (v2.0.5) and impute it using PBWT with the HRC1.1 panel (GRCh37). We excluded individuals with more than 5% missing genotypes, with too high or too low heterozygosity (inbreeding coefficient abs(F) > 0.2), duplicated measurements (keeping the one with higher quality), as well as individuals of non-European descent as determined using PCA based on ancestry informative markers. All study samples whose Euclidean distance from the center falls outside a radius of > 1.5 x maximum Euclidean distance of the European reference samples (the 1000 Genomes dataset), are considered non-European. We excluded SNPs with a call rate < 95% and actionable variants. We conducted additional quality control steps for genetic association analysis according to the guidelines^59^ using PLINK v1.90b6.24 and custom R scripts. Briefly, we checked for sex discrepancy based on X chromosome inbreeding coefficient and none had mismatches. We removed SNPs with an imputation INFO score < 0.7, minor allele frequency < 0.05, and SNPs that deviated from Hardy-Weinberg equilibrium (p < 1e-6), as well as SNPs on the sex chromosomes. We removed individuals with high or low heterozygosity rates (individuals who deviate ± 3 s.d. from the sample’s heterozygosity rate mean), resulting in a final dataset of 5,242,958 SNPs and 1,924 individuals (846 males, 1078 females).

### Genome-wide association analysis

For each protein, we adjusted rank-based inverse normal transformed levels for age, sex, BMI SDS, and plasma sample storage time. We standardized the residuals again using rank-based inverse normal transformation and used the standardized values as phenotypes and genotyping arrays as covariates for genome-wide association testing using univariate linear mixed model (LMM) implemented in GEMMA^60^ (version 0.98.5). We calculated the centered relatedness matrix to control for cryptic relatedness and population stratification. In total, 1,914 individuals passed the genotype data QC and had proteomics and covariate data available. We used the Wald test to compute all P values. Genomic inflation factor from GWAS results was calculated for each protein using the open-source Python script compute_lambda.py (version 2.0).

### Definition and refinement of significant loci

We reported the total number of identified significant associations using both a conservative multiple comparison-corrected threshold of p<1.2×10^−10^ (5×10^−8^ Bonferroni-adjusted for 420 proteins tested) and the conventional genome-wide threshold of 5×10^−8^, the latter of which was used for all downstream analysis. We defined primary pQTLs through linkage disequilibrium (LD) clumping (r^2^>=0.2) within +/- 1Mb (Plink version 1.90b6.24 with parameters --clump-r2 0.2 --clump-kb 1000) and treating the HLA region (chr6: 29691116–33054976 for hg19) as one locus, for which we report one representative SNP-protein association with the lowest P-value. We defined a cis-pQTL variant as a SNP residing within 500 Kb upstream or downstream (+/- 500 Kb) of the transcription start site (TSS) of the corresponding protein-coding gene. We extracted the TSS information for all proteins from BioMart by mapping UniProt IDs (automatic) or gene names (manual) to UniParc IDs.

Previous research has utilized various methods to report the pQTLs. These methods can be classified into two main groups: LD- and distance-based clumping or grouping ^34,35,61,62^, as well as conditional analysis aimed at identifying independent pQTLs^6–9,63–65^. In this study, we utilized the clumping approach. To account for long-range LD that may have been overlooked by this approach, we calculated the pairwise LD between all primary pQTLs for the same protein. The results showed that only a small percentage (2%) had an r^2^ greater than 0.2, indicating a low level of correlation between the primary pQTLs identified in this study. We note that the lack of unified reporting standards in pQTL studies hinders direct cross-study comparisons. Therefore, we contextualize our findings with those of previous studies by comparing the number of proteins with identified pQTLs, rather than the number of pQTLs themselves.

### Eliminating artefactual pQTLs

In MS-based proteomics, the presence of protein-altering variants can create artefactual pQTLs because the reference human proteome database used in the analysis does not contain the variant protein versions^11^. This means that after enzymatic digestion, peptides containing variant amino acids will not be quantified, and the reference version of the variant peptide can only be detected in individuals homozygous and heterozygotes for the reference allele. As a result, this creates artificial differences in the observed peptide abundances, which can bias quantification at the protein level if the reference peptide was used. To address this issue, we used a framework to assess the validity of identified pQTLs. First, all significant pQTLs (not limited to the primary pQTLs after the clumping procedure) are screened for protein-altering variants, specifically missense variants in genes encoding the same associated proteins. Next, affected peptides are then looked up in the peptide level data to see if any have been detected by MS and further used for protein quantification. Finally, a pQTL is considered an artifact if no supporting evidence from other non-affected peptides is present.

### Effect size of pQTLs

In addition to beta statistics derived from the association test, we determined the effect size of the pQTLs by calculating the ratio of mean protein abundance in heterozygotes (1/0) and homozygotes (1/1) to that of the wild type (0/0) without any data transformation.

### Comparison with previous pQTL studies

To assess the novelty of the identified pQTLs in this study, we compared our results at genome-wide significance level to 33 previously published pQTL studies. These prior studies contain more than 100,000 pQTLs above 5,000 distinct genes from various proteomics platforms dominated by the SomaScan assay and Olink panels. For all studies, we retained the pQTLs at the reported significance levels. We defined novelty of variant-protein pairs if no variants residing within +/- 1Mb of the primary pQTLs in this study have been reported in previous plasma/serum pQTL studies for the corresponding protein, and otherwise a replication. LD was not considered in this analysis. Comparison was done at protein level by matching the reported gene name from each study. Gene names are mapped based on Uniprot IDs through Uniport ID mapping in case of missing values. Genome coordinates based on GRCh38 were converted to GRCh37 using pyliftover (version 0.4).

### Mapping pQTLs to GWAS Catalogue results

We sought to identify if any of the primary pQTLs have been reported to be associated with a disease or trait. We did this by mapping the primary pQTLs to GWAS Catalogue results (v. 1.0.2) through SNP IDs. LD was not considered in this analysis.

### Replication of pQTLs in the independent cohort

The study protocol for the GALAXY replication cohorts was approved by the ethics committee for the Region of Southern Denmark (nos. S-20160006G, S-20120071, S-20160021 and S-20170087) and is registered with both the Danish Data Protection Agency (nos. 13/8204, 16/3492 and 18/22692) and Odense Patient Data Exploratory Network (under study identification nos. OP_040 and OP_239 (open.rsyd.dk/OpenProjects/da/openProjectList.jsp)). Genotype and proteomics data processing and the association analysis in the GALAXY replication cohorts of the ALD study was carried out similarly to the pipeline used for the discovery study where applicable except that 1) liver disease status (histological scoring of fibrosis F0-F4, inflammatory activity I0-5 and steatosis score S0-3), alcohol abstinent status, and statin use status were further controlled for at the protein level in addition to age, BMI and, sex and 2) genotype imputation was performed using minimac4 (version 1.0.2) and quality was filtered at R2>0.5, which has been shown to be a good threshold for separating between poorly and well-imputed variants and equivalent to an INFO score>0.7^66,67^. Out of the 1,116 primary pQTLs found in the discovery cohort, 1,037 were testable in the replication study with both variant and protein data available (n=558). A pQTL was considered replicated if the same SNP or its proxy (r^2^>0.2 within a region +/- 1Mb) is also significantly associated (p<0.05) with the protein with a concordant direction of effect. Applying a Bonferroni corrected *p-value* of 4.8×10^−5^ (0.05/1,037) led to a replication rate of 53%.

## Data Availability

The datasets produced in the present study are available upon reasonable request to the authors. All analysis results are available as supplementary tables. Searchable results are available online at proteomevariation.org

https://ftp.ebi.ac.uk/pub/databases/reference_proteomes

https://www.ebi.ac.uk/gwas/docs/file-downloads

https://grch37.ensembl.org/info/data/biomart/index.html

## Data availability

The human reference proteome database (2018 release, both canonical and additional sequences) was downloaded from the European Bioinformatics Institute database (https://ftp.ebi.ac.uk/pub/databases/reference_proteomes/); The GWAS Catalog (v.1.0.2) was downloaded at https://www.ebi.ac.uk/gwas/docs/file-downloads; Transcription start site of proteins was extracted from BioMart accessed in August 2022 (https://grch37.ensembl.org/info/data/biomart/index.html); The datasets generated and/or analysed during the current study are not publicly available due to need to maintain privacy of study participants, but are available from the corresponding authors on reasonable request. All analysis results are available as supplementary tables. Searchable results are available online at proteomevariation.org. The study protocol is also available upon request to Jens-Christian Holm.

## Code availability

The software used in this study can be accessed here: Spectronaut (v15.4): https://biognosys.com/software/spectronaut/; Python (v3.9.0 and 3.8.11): https://www.python.org/; Combat (v0.3.2): https://pypi.org/project/combat/; Perseus (v1.6.5.0): https://maxquant.net/perseus/; Compute-lambda.py (v0.2): https://github.com/pgxcentre/lambda; bcftools (v1.14): https://samtools.github.io/bcftools/; Gemma (v0.98.5): https://github.com/genetics-statistics/GEMMA; Plink (v1.90b6.24): https://www.cog-genomics.org/plink/; Variant effect predictor was performed online with the RefSeq transcript database (http://grch37.ensembl.org/Homo_sapiens/Tools/VEP); Scikit-learn (v1.0): https://scikit-learn.org/stable/whats_new/v1.0.html; INT-transformation: https://github.com/edm1/rank-based-INT; Scipy (v1.7.1): https://scipy.org/; Pingouin (v0.4.0): https://pingouin-stats.org/build/html/index.html; Pyliftover (v0.4): https://pypi.org/project/pyliftover/; The customed scripts can be accessed at github.com/llniu/pQTL_HolbaekStudy.

## Acknowledgements

Our gratitude goes to all participants and their families from The HOLBAEK Study. We appreciate the staff at The Children’s Obesity Clinic for their assistance in clinical studies, especially Gitte Holløse and Tanja Larsen for providing state of the acquisition, storage, and maintenance of biological samples and clinical data. Jette Bork Jensen and Jesus Vicente Torresano Lominchar at the Phenomics Platform of Novo Nordisk Foundation Center for Basic Metabolic Research deserve recognition for their genomics data processing contributions. We also acknowledge the members of the Clinical Proteomics Group at Novo Nordisk Foundation Center for Protein Research and the Department of Proteomics and Signal Transduction (Max Planck Institute of Biochemistry). Special thanks to Lylia Drici and Andreas Brunner for their technical assistance. Jeppe Madsen and Michael Wierer, Director of Proteomics Research Infrastructure (PRI), provided valuable technical support as well. Our appreciation extends to Valborg Gudmundsdottir at the University of Iceland for kindly sharing pQTL results she had collected from existing 20 pQTL studies and Lasse Folkersen at Nucleus for discussions. Finally, we acknowledge the funding agencies that supported this study: the Novo Nordisk Foundation for the Clinical Proteomics Group (grant no. NNF15CC0001 to M.M.), the Challenge Program (grant no. NNF15OC0016692 to the MicrobLiver consortium), the Innovation Fund Denmark (grant no. 0603-00484B to T.H.), and the Novo Nordisk Foundation (grant no. NNF15OC0016544), while the European Union’s Horizon 2020 research and innovation program (grant no. 668031 to the GALAXY consortium) funded the GALAXY cohorts in the ALD study. Additionally, L.N. and S.R. received support from the Novo Nordisk Foundation (grant no. NNF14CC0001 and NNF21SA0072102 to S.R.), M.T. from the Novo Nordisk Foundation (grant no. NNF20OC0059393), C.E.F. received support from the BRIDGE – Translational Excellence Program (grant no. NNF18SA0034956) and the Region Zealand Health and Medical Research Foundation (grant no. R32-A1191).

## Contributions

M.M., T.H., L.N. and S.S. conceived the project and designed the experiments. L.N., performed the proteomics experiment, led the computational analyses, interpreted the data, generated the figures, and wrote the manuscript. S.S. selected the samples to be included in this study, contributed to designing the computational pipeline, and revised the manuscript. L.A.H., M.A.V.L., C.E.F., and J.H contributed with the clinical data and revised the manuscript. L.C., J.M. contributed to designing the computational pipeline and revised the manuscript. H.B.J. revised the manuscript. M.T. and A.K. contributed with clinical data in the validation cohort. S.R. oversaw the computational pipeline for data analysis and revised the manuscript. T.H. contributed with the genomics data and revised the manuscript. L.N. and M.M. wrote and edited the manuscript with input from all coauthors.

## Materials & Correspondence

Correspondence and material requests should be addressed to Simon Rasmussen, Torben Hansen and Matthias Mann (Lead contact).

## Competing interests

M.M. is an indirect investor in Evosep.

## Notes

### Funding Statement

This study was funded by the Novo Nordisk Foundation for the Clinical Proteomics Group (grant no. NNF15CC0001 to M.M.), the Challenge Program (grant no. NNF15OC0016692 to the MicrobLiver consortium), the Innovation Fund Denmark (grant no. 0603-00484B to T.H.), and the Novo Nordisk Foundation (grant no. NNF15OC0016544), while the European Union Horizon 2020 research and innovation program (grant no. 668031 to the GALAXY consortium) funded the GALAXY cohorts in the ALD study. Additionally, L.N. and S.R. received support from the Novo Nordisk Foundation (grant no. NNF14CC0001 and NNF21SA0072102 to S.R.), M.T. from the Novo Nordisk Foundation (grant no. NNF20OC0059393), C.E.F. received support from the BRIDGE - Translational Excellence Program (grant no. NNF18SA0034956) and the Region Zealand Health and Medical Research Foundation (grant no. R32-A1191).

### Author Declarations

Ethics committee for the Region Zealand and Region of Southern Denmark in Denmark gave ethical approval for this work.

