## Supplemental Table 1 for "Plasma Proteome Variation and its Genetic Determinants in Children and Adolescents"

Supplementary Table 1: Participant characteristics in the discovery and replication cohorts

| | Discovery cohort | | Replication cohort |
|---|---|---|---|
| | General population (n=951) | Obesity clinic (n=1179) | GALA-ALD/HP (n=558) |
| Age, mean (s.d.), y | 12 (3) | 12 (3) | 56 (10) |
| Female sex, No. (%) | 530 (56) | 641 (54) | 148 (73) |
| Male sex, No. (%) | 421 (44) | 538 (46) | 410 (27) |
| BMI, mean (s.d.), kg/m2 | 17.7 (2.3) | 27.1 (5.6) | 27.2 (5) |
| BMI SDS, mean (s.d.) | -0.05 (0.81) | 2.77 (0.75) | NA |
| Tanner stage 1/2-5, No. (%) | 233/474 (33/67) | 327/569 (36/64) | NA |
| ALT, median (IQR), U/L | 19 (16-23) | 24 (19-31) | 28 (21-42) |
| AST, median (IQR), U/L | 26 (22-31) | 24 (20-29) | 30 (24-46) |
| GGT, median (IQR), U/L | 16 (12-19) | 17 (15-21) | 48 (25-136) |
| Glucose, median (IQR), mmol/L | 5 (4.7-5.2) | 5 (4.8-5.3) | 6 (5.5-6.6) |
| Insulin, median (IQR), pmol/L | 51 (36.3-68.4) | 79 (55.1-117.8) | NA |
| HbA1c, median (IQR), mmol/mol | 34 (32-35) | 34 (32-36) | 36 (33-39) |
| Triglycerides, median (IQR), mmol/L | 0.6 (0.5-0.8) | 0.9 (0.7-1.3) | 1.2 (0.9-1.8) |
| Total cholesterol, median (IQR), mmol/L | 3.9 (3.5-4.3) | 4 (3.6-4.6) | 5 (4.4-5.9) |
| LDL cholesterol, median (IQR), mmol/L | 2 (1.7-2.4) | 2.3 (1.9-2.8) | 3 (2.3-3.6) |
| HDL cholesterol, median (iQR), mmol/L | 1.5 (1.3-1.8) | 1.2 (1-1.4) | 1.3 (1.1-1.7) |
| Abstaining from alcohol at time of inclusion, No. (%) | NA | NA | 197 (35) |
| Statin use prior time of inclusion, No. (%) | NA | NA | 97 (17) |
| Steatosis 0/1/2/3, No. | NA | NA | 373/79/70/36 |
| Inflammatory activity 0/1/2/3/4/5, No. | NA | NA | 290/90/78/50/28/22 |
| Fibrosis stage 0-1/2/3/4, No. | NA | NA | 367/102/26/63 |

BMI=body mass index; SDS=standard deviation score; IQR= Interquartile range; ALT=alanine aminotransferase; AST=aspartate aminotransferase; GGT=gamma glutamyl transferase; HbA1C=hemoglobin A1c; LDL=low-density lipoprotein; HDL=high-density lipoprotein.
